# The Presence of a Paramagnetic Phase Rim is Linked to Lesion Age in Multiple Sclerosis

**DOI:** 10.1101/2025.01.12.25320426

**Authors:** Samuel Klistorner, Tatiana Usnich, Margareta A. Clarke, Deborah Pareto, Àlex Rovira, Friedemann Paul, Michael Barnett, Alexander Klistorner

**Author notes:** These authors contributed equally to this work (Samuel Klistorner, Alexander Klistorner). Correspondence to: Alexander Klistorner, Full address: Save Sight Institute, 8 Macquarie St, Sydney NSW, Australia, postcode 2000.

## Abstract

**Background and Objectives:** In relapsing-remitting multiple sclerosis (RRMS), smouldering inflammation at the rims of chronic active lesions has emerged as a crucial contributor to disease progression. Paramagnetic rim lesions (PRLs), has been proposed as the most pathologically validated marker for chronic active lesions. However, the strength of the association between PRLs and clinical or radiological measures of disease progression remains unclear, and the relationship between PRL presence and lesion-specific characteristics, such as lesion size and age, is not well understood. The objective of this study is to investigate the effect of PRL presence on clinical and radiological markers of disease progression, and its association with lesion characteristics such as size and age.

**Methods:** 60 RRMS patients, each with at least five years of previous structural MRI data were investigated using SWAN protocols. Lesions exceeding a volume of 100 mm³ were included in analysis.

**Results:** PRLs were present in 48% of patients, comprising 13% (80 out of 607) of the total lesion count. Patients with PRLs were significantly younger than those without (p<0.001). Furthermore, PRLs were significantly larger (p<0.0001) and exhibited more severe structural damage compared to non-PRLs (p<0.0001). These characteristics were consistent both within and between patients. PRL count and volume were significantly correlated with radiological measures of disease progression, including central and total brain atrophy (p<0.001 and <0.05 for PRL count and volume, respectively).

Crucially, our results showed that all 32 lesions appearing within five years preceding SWAN imaging displayed a paramagnetic rim. This finding was validated in two independent international cohorts, reinforcing the link between PRLs and lesion age. Moreover, the proportion of rim-positive lesions decreased as lesion age increased. In a sub-set of patients with longitudinal susceptibility data the paramagnetic rim tended to diminish or disappear over time.

**Conclusion:** Our findings indicate that the presence of a paramagnetic rim in MS lesions is a characteristic feature of all newly formed lesions that exceed a specific size threshold, and that this rim gradually diminishes as lesions age. As such, this study enhances the understanding of lesion formation and may also have significant implications for using PRLs as a biomarker of lesion activity.

## Introduction

Multiple sclerosis (MS) is a complex central nervous system (CNS) disorder characterised by heterogeneous inflammatory and variably destructive white and grey matter lesions.

Among the various pathological features of MS, smouldering inflammation—particularly at the rims of chronic active lesions—has emerged as a crucial driver of disease progression. Histopathologically, these chronic active lesions (CAL), also known as "smouldering" lesions, exhibit an inactive, demyelinated core surrounded by a narrow rim of activated macrophages/microglia that contain myelin degradation products, indicative of ongoing low-grade demyelination.^12^ This smouldering inflammation around CAL leads to slow but persistent axonal injury, contributing to progressive neurodegeneration, loss of brain tissue and disability progression.^3–6^

Despite its importance, the nature of smouldering inflammation and the methods to accurately identify it remain poorly understood. This lack of clarity contributes to its frequent under-detection, presenting a major challenge in the effective management of MS. ^35^ Consequently, smouldering inflammation has only recently become a primary target for emerging therapeutics, such as Bruton’s tyrosine kinase inhibitors (BTKi). While early data suggests that this approach may translate to clinical benefit in the form of reduced disability worsening,^7^ smouldering inflammation remains an unmet therapeutic target for people living with MS, underscoring the need for effective biomarkers to monitor and understand this phenomenon.

Several promising imaging biomarkers for CAL have been identified,^8^ including paramagnetic rim lesions (PRLs) on susceptibility-sensitive MRI sequences, slow expansion of chronic lesions on T1 or T2-weighted images, and 18-kDa translocator protein (TSPO) - positive lesions on PET. Among these biomarkers, PRLs, a subset of chronic focal white matter lesions in which the peripheral rim is visualized on MRI using susceptibility/T2*-weighted sequences, has been proposed as the most pathologically validated marker for chronic inflammation. ^8–11^

However, while the relationship between the presence of PRL and clinical and radiological measures of disease progression has been widely reported in recent years,^12–22^ the strength of this association remains unclear.

The temporal dynamics of PRLs also remain uncertain. Although PRLs have been observed to arise at the time of lesion formation,^8^ the reported proportion of new lesions developing a paramagnetic rim varies considerably, ranging from 0 to over 50%. ^4,12,23–27^ Furthermore, recent studies have demonstrated a significant discrepancy between the presence of a paramagnetic rim and slow lesion expansion, another biomarker of chronic active lesions.^28,29^

Given these uncertainties, the objective of this study is to investigate the effect of PRL presence on clinical and radiological markers of disease progression, and its association with lesion characteristics such as size and age. This analysis seeks to clarify the role of PRLs in understanding MS pathology and monitoring smouldering inflammation. To enhance the reliability of our findings, the results of the primary analysis were validated using two independent patient cohorts.

## Materials and methods

The study was approved by the University of Sydney Human Research Ethics Committees and followed the tenets of the Declaration of Helsinki. Written informed consent was obtained from all participants.

### Subjects and study design

Consecutive patients with established relapsing-remitting MS (RRMS; diagnosed according to the revised McDonald 2010 criteria^30^) who were enrolled in Mechanism of Axonal Degeneration in MS study, had reached at least 60 months of follow-up, underwent annual MRI on the same 3 T MRI system and had susceptibility-weighted angiography (SWAN) imaging at the last time point (referred to as SWAN timepoint) were included (Fig 1).

**Figure 1.**
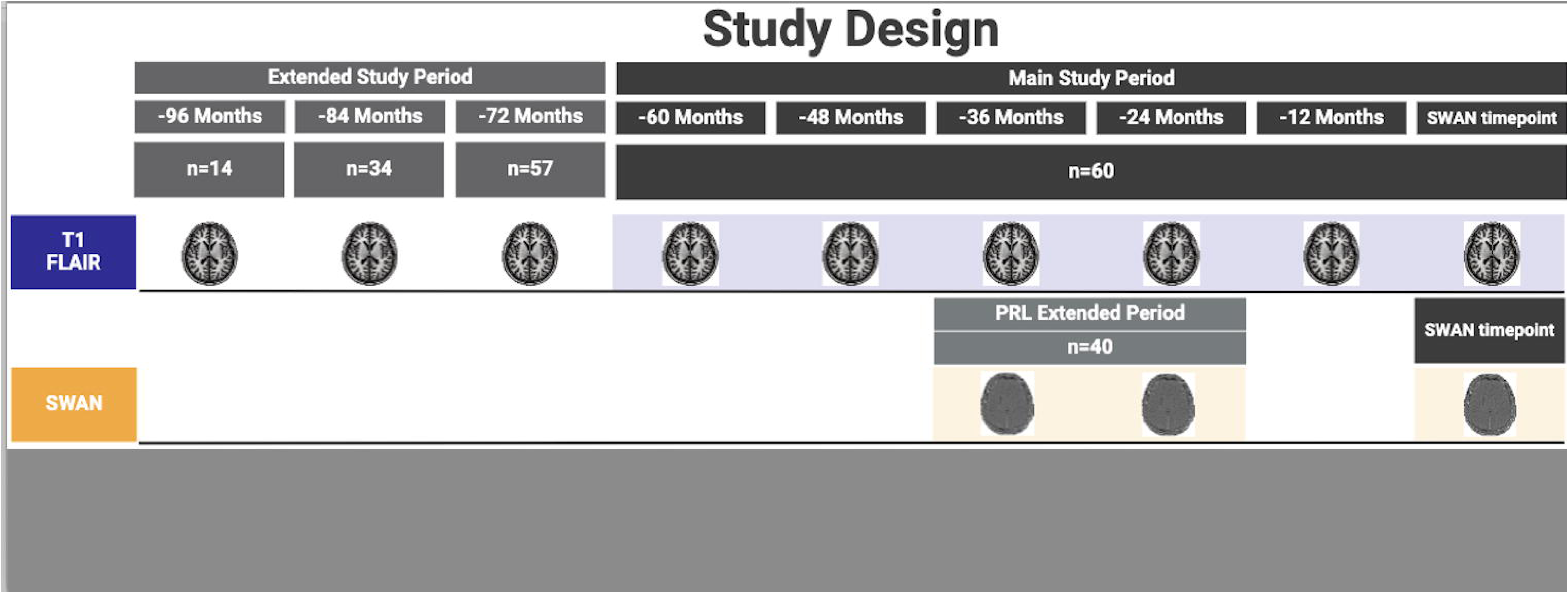
Schematic representation of the study design. This figure provides an overview of the study’s structure, including key time points, imaging intervals, and assessment phases used to track PRL changes over the extended study period.

The SWAN timepoint was used for all cross-sectional analyses; and the data obtained within the 60 months prior (referred to as the main study period) was used for the longitudinal analysis.

The extended study period (up to 96 months prior to the SWAN timepoint), with a reduced number of patients, was used for a sub-study analysis.

### MRI protocol

MRI was performed using a 3T GE Discovery MR750 scanner (GE Medical Systems, Milwaukee, WI). The following MRI sequences were acquired: pre- and post-contrast (gadolinium) sagittal 3D T1-weighted, 3D T2-FLAIR, diffusion-weighted imaging and SWAN protocol using a 3D gradient-echo sequence. Specific acquisition parameters and MRI image processing are presented in the Supplementary material.

### Lesion analysis

White matter lesions were segmented for each time point of each patient using an advanced AI-based lesion segmentation algorithm (iQ-Solutions™ MS Report, Sydney Neuroimaging Analysis Centre, Sydney, Australia). For this analysis, only lesions exceeding a volume of 100 mm³ were included to maintain consistency and ensure the paramagnetic rim could be clearly visualized on phase imaging.

The degree of tissue damage within these chronic lesions was quantified using T1 hypointensity and mean diffusivity (MD), as described in previous studies^31^ and specified in the Supplementary material.

### PRL analysis

For each patient, PRLs were visually identified on the phase images at SWAN timepoint scan according to criteria proposed in the 2022 NAIMS Consensus Statement on Imaging Chronic Active Lesions^8^. The senior author (AK) assessed the presence of rim lesions.

### Volumetric brain analysis

Total brain, white matter, grey matter and ventricular volume^32,33^ were analysed using AssemblyNet, an AI brain segmentation tool, on T1 images co-registered to ACPC space.^34^ All volumes were scaled based on MNI template registration to account for head size variability and as such, the measures at the SWAN timepoint are reported as a % of total intracranial volume (% of TIV). For longitudinal analysis, the annualised percentage change during the main study period (i.e. 5 years) was calculated for each MRI measure to assess atrophy.

### Validation datasets

To validate the relationship between the presence of a paramagnetic rim and lesion age, a consecutive cohort of RRMS patients from Charité Universitaetsmedizin Berlin in Germany and RRMS patients and clinically isolated syndrome (CIS) form Vall d’Hebron University Hospital in Barcelona, Spain were included. Each patient underwent at least 3 consecutive MRI scans. Similar to the main study, PRLs were assessed at the final time point. Detailed information on the study protocols from the two cohorts have been published previously^35,36^ Imaging protocols are listed in the Supplementary Table 1.

### Statistics

Continuous variables were assessed for normality using the Shapiro–Wilk test and described as either the mean with standard deviation (SD) or median with interquartile range (IQR), depending on the distribution.

For group comparisons between subjects with and without PRLs, the Chi-square test was used to compare sex distributions and baseline Disease-Modifying Therapy (DMT) usage. Additionally, Fisher’s Exact Test was employed to evaluate the relationship between PRL presence and changes in treatment lines between groups. Differences in cross-sectional Expanded Disability Status Scale (EDSS) scores between groups were assessed using the Independent-Samples Mann-Whitney U Test, while longitudinal changes in EDSS were evaluated with the Related-Samples Wilcoxon Signed Rank Test. Age and disease duration differences between groups were analysed using the Student’s t-test.

An Analysis of Covariance (ANCOVA) was conducted to determine significant differences in brain atrophy measures between groups, with cross-sectional and longitudinal analyses adjusted for age. For individual lesion analysis, T1 intensity and MD values were adjusted for lesion volume using ANCOVA.

Spearman’s Rank Correlation was applied to assess the relationship between the count or volume of PRLs and various clinical and imaging metrics, including EDSS scores, brain volume, and atrophy rates.

As this was an exploratory study, no adjustment for multiple comparisons was performed.

## Results

The main study cohort, MADMS, included sixty consecutive MS patients enrolled in a longitudinal observational study, all of whom had reached at least 60 months of follow-up and had one SWAN scan at the latest follow-up.

At the SWAN timepoint, the average age of the cohort was 46.9 ± 9.9 years, with 44 females (73%) and 16 males. The patients had a mean disease duration of 13.8 ± 6.0 years, and the median EDSS was 1.0 (interquartile range: 2). Disease-modifying therapy categories for main cohort described in the Supplementary material.

Of all the patients, 29 (48%) exhibited at least one PRL. In total, 80 out of 607 lesions were identified as PRLs, representing 13% of the total lesion count.

### Group Comparison Between Patients With and Without PRL (Patient-Based Analysis)

A comparison between PRL-positive and PRL-negative subjects is presented in Table 1. Patients with PRL were significantly younger (p<0.001). Both groups had an equal male-to-female ratio and similar total lesion volume, total lesion count and disease duration.

**Table 1.** Demographic and volumetric data between PRL(+) and PRL(-) subjects.

| Characteristic | PRL(+)<br>Subjects | PRL(-) Subjects | p-value |
| --- | --- | --- | --- |
| Number | 29 | 31 |  |
| **Demographics** |  |  |  |
| Age, mean (Std) | 42.31 (8.39) | 51.1 (9.29) | **<0.001** |
| Sex (m/f, %m) | 8/21 (28%) | 8/23 (26%) | 0.891 |
| Disease duration, mean (Std) | 11.62 (5.48) | 14.0 (6.28) | 0.135 |
| **Lesion** |  |  |  |
| Lesion Volume, mean (Std) | 7823 (8611)<br>mm<sup>3</sup> | 5719 (7225)<br>mm<sup>3</sup> | 0.317 |
| Lesion Count, mean (Std) | 10.59 (8.22) | 9.4 (7.77) | 0.581 |
| **EDSS** |  |  |  |
| SWAN timepoint EDSS (median, IQR) | 1.0 (0.0 – 2.0) | 1.0 (1.0 – 2.0) | 0.078 |
| EDSS change (median, IQR) | 0.0 (0.0 – 0.5) | 0.0 (0.0 – 0.5) | 0.633 |
| SWAN timepoint measures (% of TIV) |  |  | p-value adj. for age |
| Ventricle, mean (Std) | 1.79 (0.65) | 1.9 (0.86) | 0.870 |
| White matter, mean (Std) | 28.02 (2.06) | 28.6 (1.88) | **0.014** |
| Grey matter, mean (Std) | 45.43 (1.04) | 45.0 (1.13) | 0.987 |
| Parenchymal brain, mean (Std) | 84.78 (3.13) | 84.9 (2.98) | **0.049** |
| Atrophy (annualised % change) |  |  | p-value adj. for age |
| Ventricle, mean (Std) | 1.80 (2.04) | 2.0 (2.45) | 0.323 |
| White matter, mean (Std) | -0.39 (0.34) | -0.4 (0.41) | 0.078 |
| Grey matter, mean (Std) | -0.32 (0.16) | -0.2 (0.13) | 0.115 |
| Total brain, mean (Std) | -0.32 (0.19) | -0.3 (0.18) | 0.057 |

When adjusting for age, PRL-positive patients exhibited a smaller intracranial fraction of parenchymal brain (PBF) (Table 1) (p=0.049) and white matter (p=0.014) at the SWAN timepoint. Longitudinal analysis, also adjusted for age, revealed a small difference in total brain atrophy of borderline significance (PRL-positive vs. PRL-negative, p=0.05). However, there was no significant difference between the two groups in the central brain atrophy (ventricular volume), white matter atrophy or grey matter atrophy.

No significant differences were observed in cross-sectional EDSS measures or longitudinal EDSS changes between the groups. The distribution of cross-sectional DMT categories/lines was similar between the groups (χ2=0.82, p=0.66). Additionally, treatment advancement between categories/lines remained non-significant between the two groups (Fisher’s statistic = 2.97, p=0.17).

### Group comparison between PRL and non-PRL lesions (Lesion based analysis)

The relationship between PRLs and non-PRLs across various metrics is shown in Table 2.

**Table 2:** Lesion comparison between PRL and non-PRL for entire cohort and PRL+ patients only.

|  | All PRLs | Non-PRLs in Entire cohort (60 subjects) | p-value | Non-PRLs In PRL+ subjects only (29 subjects) | p-value |
| --- | --- | --- | --- | --- | --- |
| Lesion count | 80 | 527 |  | 262 |  |
| Lesion volume, mean (Std) | 1509 (2672) mm<sup>3</sup> | 499 (1008) mm<sup>3</sup> | **<0.0001** | 451 (927) mm<sup>3</sup> | **<0.0001** |
| Lesion T1 intensity value, mean (std) | -15.0 (5.93) | -12.1 (5.64) | **<0.0001** | -11.9 (5.53) | **<0.0001** |
| Lesion MD, mean (std) | 1.18 (0.15) μm<sup>2</sup>/ms | 1.08 (0.14) μm<sup>2</sup>/ms | **<0.0001** | 1.08 (0.14) μm<sup>2</sup>/ms | **<0.0001** |

The combined analysis of all 607 lesions (80 PRL-positive and 527 PRL-negative lesions) across the entire cohort of 60 patients revealed that PRL-positive lesions had a significantly larger average volume (p<0.0001). Additionally, the degree of tissue damage was more severe in PRL-positive lesions, as indicated by T1 intensity values and MD measurements (p<0.0001 for both).

These differences were also observed in an intra-subject comparison performed using 29 patients who had both PRL-positive and PRL-negative lesions (80 PRL-positive and 262 PRL-negative lesions). Within this subgroup, PRL-positive lesions were also significantly larger than non-PRL lesions (p<0.0001, paired t-test), and exhibited more severe tissue damage within the same individual (p<0.0001 for both).

### Relationship Between PRL Count and Volume vs Disease Progression

The relationship between PRL count and volume versus various disease progression metrics is shown in Table 3. PRL count demonstrated a strong positive correlation with total lesion volume (ρ = 0.550, p = 0.002) and total lesion number (ρ = 0.572, p = 0.001). Similarly, PRL volume showed significant correlations with total lesion volume (ρ = 0.801, p < 0.001) and total lesion number (ρ = 0.668, p < 0.001). In addition, PRL volume showed borderline correlation with disease duration (ρ = 0.375, p = 0.049).

**Table 3:** Correlation between PRL count or PRL volume and demographic and radiological measures.

|  | PRL Count |  | PRL Volume |  |
| --- | --- | --- | --- | --- |
| Analysis | $\rho$ (Spearman) | p-value | $\rho$ (Spearman) | p-value |
| Age | 0.137 | 0.486 | 0.152 | 0.441 |
| Disease duration | 0.17 | 0.388 | **0.375** | **0.049** |
| Lesion Volume | **0.55** | **0.002** | **0.801** | **<0.001** |
| Lesion number | **0.572** | **0.001** | **0.668** | **<0.001** |
| SWAN timepoint EDSS | 0.284 | 0.143 | 0.128 | 0.517 |
| EDSS change | 0.309 | 0.117 | **0.473** | **0.013** |
| Volumetric measures at SWAN timepoint (% of TIV) |  |  |  |  |
| Ventricle | **0.394** | **0.038** | 0.348 | 0.070 |
| White matter | **-0.519** | **0.005** | **-0.681** | **<0.001** |
| Grey matter | -0.120 | 0.543 | -0.210 | 0.284 |
| Parenchymal brain | -0.362 | 0.059 | **-0.531** | **0.004** |
| Annualised atrophy (% change) |  |  |  |  |
| Ventricle | **0.531** | **0.004** | **0.397** | **0.036** |
| White matter | **-0.414** | **0.029** | -0.305 | 0.114 |
| Grey matter | -0.325 | 0.091 | -0.268 | 0.168 |
| Total brain | **-0.486** | **0.009** | **-0.404** | **0.033** |

There was no statistically significant association between PRL count and baseline EDSS or EDSS change. However, PRL volume showed a significant positive correlation with EDSS change (ρ = 0.473, p = 0.013).

At the SWAN timepoint, the PRL count correlated significantly with white matter volume (ρ = -0.519, p = 0.005), and ventricle volume (ρ = 0.394, p = 0.038). Longitudinal atrophy metrics further supported these findings. PRL count at the SWAN timepoint significantly correlated with total brain atrophy (ρ = -0.486, p = 0.009), ventricle volume change (ρ = 0.531, p = 0.004) and white matter volume change (ρ = -0.414, p = 0.029).

Similarly, the PRL volume at the SWAN timepoint correlated significantly with disease duration (ρ = 0.375, p = 0.049), total brain volume (ρ = -0.531, p = 0.004) and white matter volume (ρ = -0.681, p < 0.001). Longitudinal atrophy metrics also demonstrated significant correlation of PRL volume at the SWAN timepoint with total brain volume (ρ = -0.404, p = 0.033) and ventricle volume change (ρ = 0.397, p = 0.036).

### PRL and Lesion Age

Thirteen out of 60 patients developed 32 new lesions within susceptibility visible areas (i.e. areas free of potential susceptibility distortion) during the main study period of 60 months. Remarkably, all 32 of these new lesions exhibited a paramagnetic rim. Examples are shown in Fig. 2.

**Figure 2:**
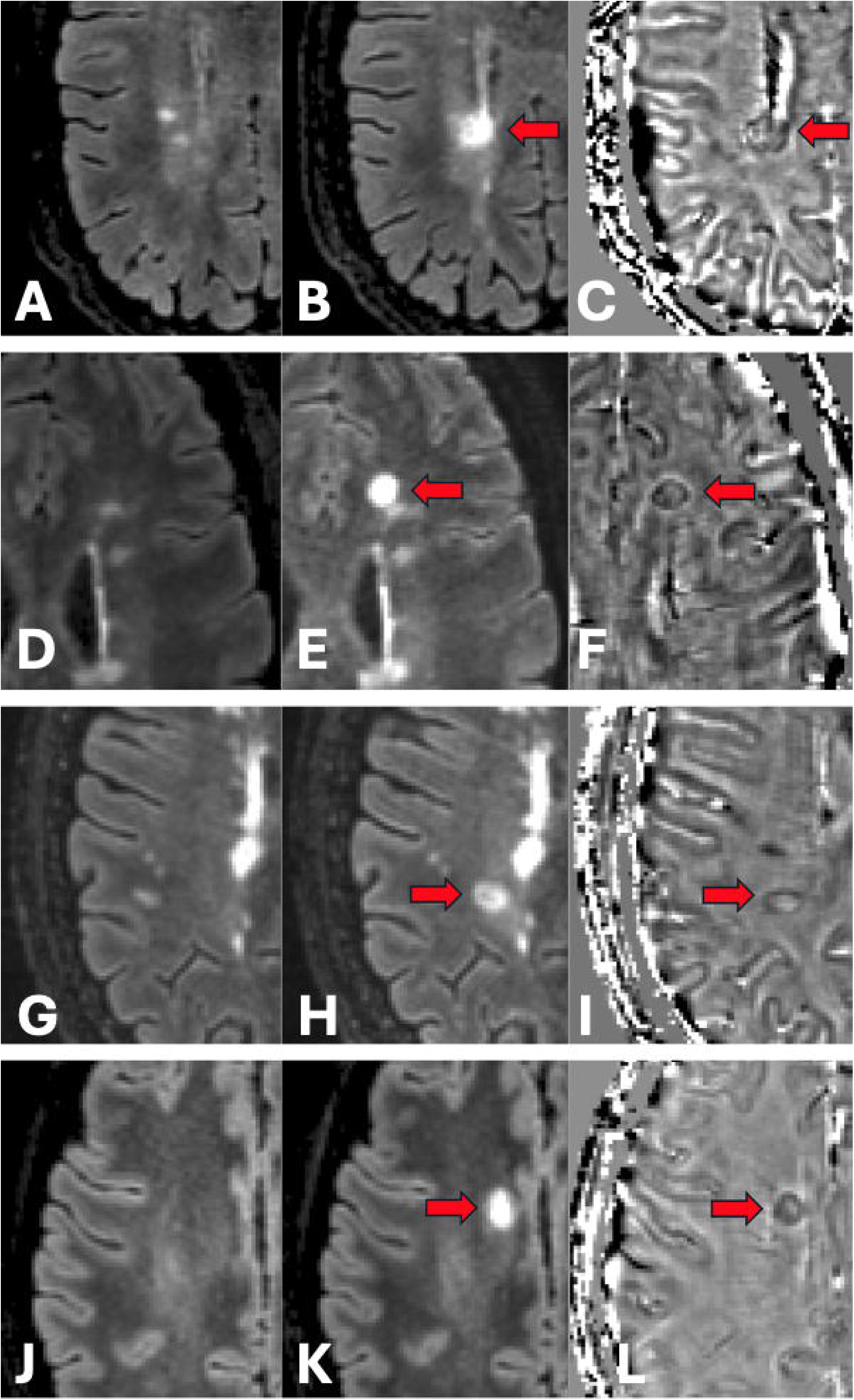
Examples of new on-study lesions and rim at SWAN timepoint. Each row represents an individual patient. **Column 1 (A, D, G, J):** FLAIR image at baseline time points. **Column 2 (B, E, H, K):** FLAIR image of new lesion appearance at SWAN timepoint and **Column 3 (C, F, I, L):** Phase image at SWAN timepoint.

In five of these 13 patients, SWAN scans were available at the initial appearance of these lesions (for instance, at the “-36 months” time point). In every case, the paramagnetic rim was already visible at that early stage of lesion formation and remained detectable up to the SWAN timepoint. Additionally, among the 13 patients, 2 had no lesions that were older than 60 months (i.e. all lesions appeared during 5-year follow-up period). In 6 of the remaining patients, although they had lesions older than 60 months, none of these older lesions displayed a paramagnetic rim. However, five patients also exhibited PRLs in lesions older than 60 months (i.e. in lesions already present at “-60 months” time point).

This is illustrated in Fig 3a, which shows the proportion of non-rim (green), new on-study rim-positive (red), and pre-study rim-positive (blue) lesions in those patients. As depicted in Fig 3b, the age of on-study rim-positive lesions was distributed across the entire follow-up period.

**Figure 3:**
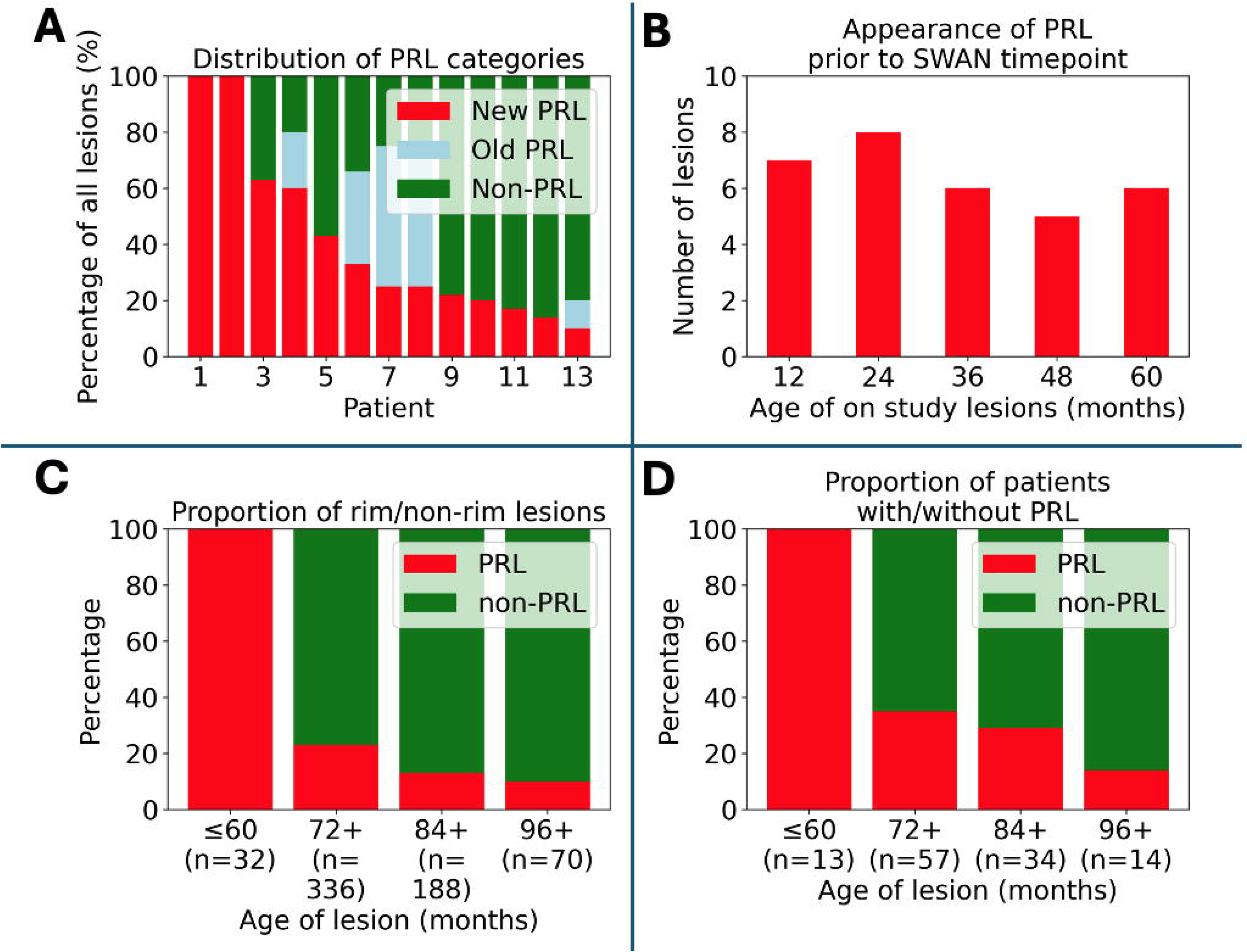
Analysis of paramagnetic rim lesions (PRLs) and non-PRLs proportions and distribution in patients. (A) Distribution of non-PRL, old PRL (which appeared before the 60-month timepoint), and new on-study PRL lesions across 13 patients with new on-study lesions. (B) Distribution of the appearance timing of the 32 on-study lesions prior to the SWAN timepoint. (C) The proportion of lesions with a rim during the extended study period. (D) The proportion of patients with PRLs, grouped by lesion age categories.

As stated in the Methods section, some patients had conventional MRI data available from periods exceeding 60 months (72 to 96 months prior to the SWAP time point). When analysing data from PRL-positive patients over the extended study period, it became evident that the proportion of rim-positive lesions decreased as the minimum lesion age increased. While all lesions younger than 60 months (i.e. on-study lesions) had a paramagnetic rim, only 23% of lesions age 72 months or more were rim-positive. This percentage dropped further to 13% for lesions at least 84 months old and 10% for lesions at least 96 months old (Fig. 3c). A similar trend was noted at the patient level: all (i.e. 100%) patients with lesions less than 60 months old demonstrated PRL, while only 35% of patients with lesions older than 72 months had a PRL, decreasing to 29% at 84 months and 14% at 96 months (Fig. 3d).

It should be noted that lesions labelled as "at least 72 months old" include all lesions visible on scans 72 months prior, meaning some lesions are likely to be older than 72 months. Similar reasoning also applies to lesions at 84 and 96 months.

### Rim decay

Out of the 60 patients, 40 had susceptibility imaging available at an additional time point, 24– 36 months before the SWAN timepoint. Among these 40 patients, 24 had at least one PRL in the earlier scans, with a total of 61 PRLs identified. In analysing longitudinal changes, 13 patients (with 16 lesions in total) showed a clear reduction or disappearance of the rim over time (Fig. 4).

**Figure 4:**
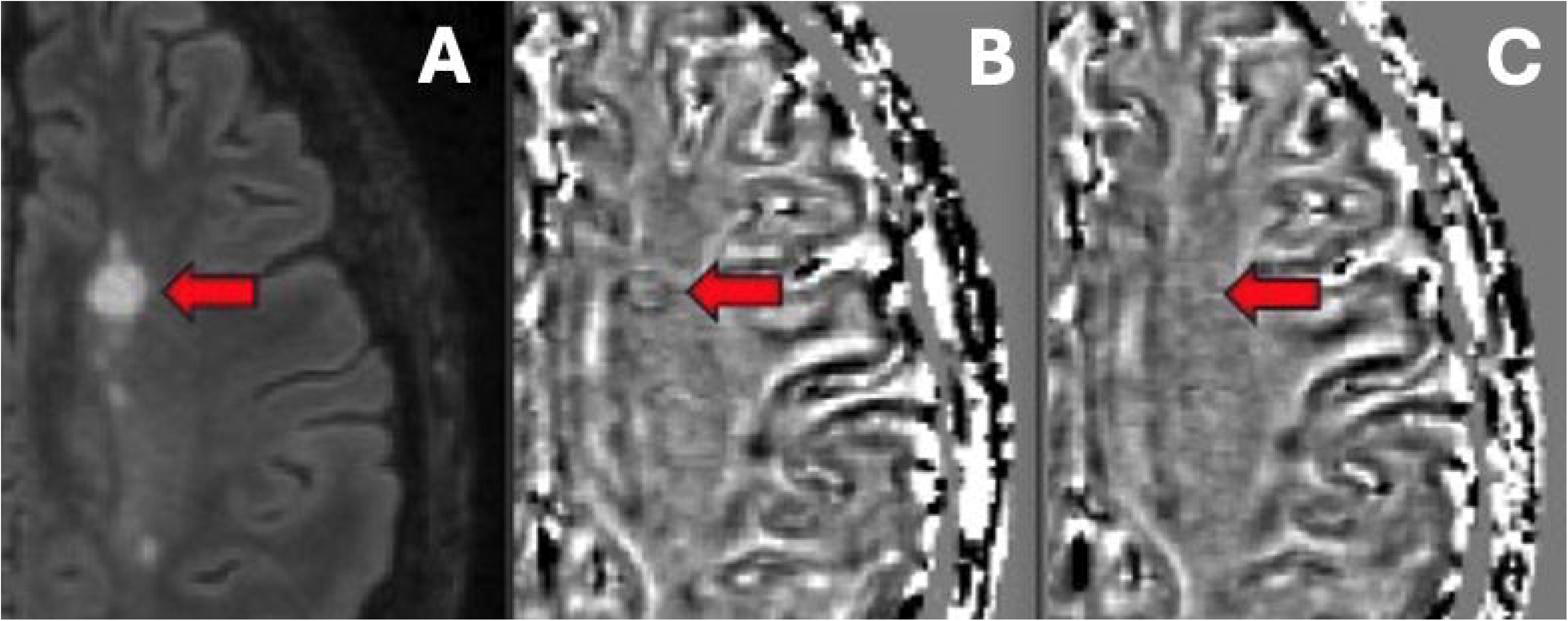
An example of rim decay. (A) Lesion at SWAN timepoint. (B) PRL on phase image 36 months prior to SWAN timepoint. (C) No clear rim at the SWAN timepoint.

### Validation of association between the presence of paramagnetic rim and lesion age

To validate the relationship between the presence of a paramagnetic rim and lesion age, data from additional cohorts were used, including 100 RRMS patients from Charité-Universitätsmedizin Berlin, Germany, and 163 CIS and MS patients from Vall d’Hebron University Hospital, Barcelona, Spain. The average time of follow-up was 43 ±23 months for the Berlin cohort and 43 ± 19 months for the Barcelona cohort.

Twelve patients from Berlin cohort developed a total of 20 new lesions during the study, an example of a PRL is illustrated in Figure 5. In the Barcelona cohort, seven patients developed new lesions but only three patients had new lesions that met the size/location criteria for a total of five new, eligible lesions. Notably, all new lesions that met the size and location criteria from both the Berlin and Barcelona cohorts exhibited a paramagnetic rim on phase image.

**Figure 5:**
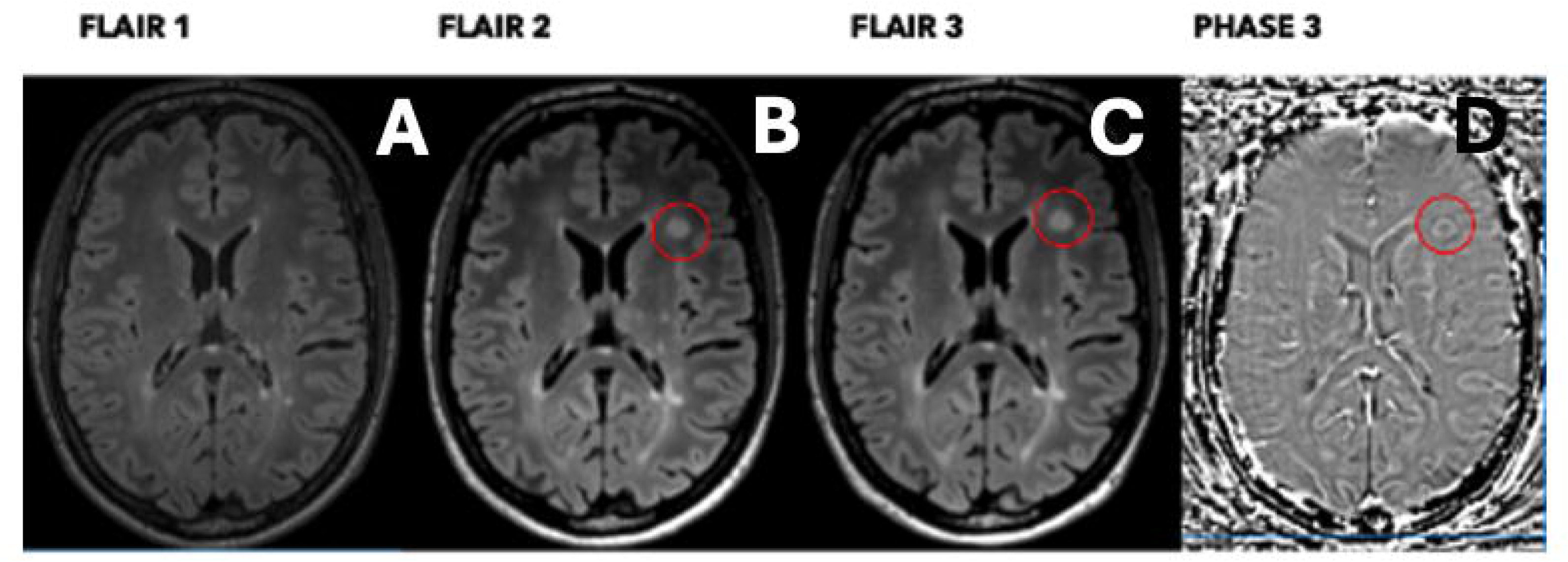
An example of on-study new lesion validation. FLAIR images at the (A) baseline time point, (B) intermediate time point and (C) last time point. Phase image (D) at the last time point. The circle highlights a new on-study lesion.

## Discussion

This study investigated the incidence of PRLs, their relationship with lesion characteristics such as lesion size and age, and their association with clinical and radiological markers of disease progression in a substantial cohort of RRMS patients. PRLs were detected in nearly half of the study population, with a higher prevalence in younger patients. These rim-positive lesions were notably larger than their non-PRL counterparts. Importantly, we demonstrated a strong negative association between the presence of a paramagnetic rim and lesion age, a finding that was further validated in two independent cohorts.

### Incidence of PRL

Our observation that PRLs were present in 48% of patients in the main study cohort aligns with previous studies, which reported PRL in 35-63% of MS patients.^12,14,15,17,19–22,37,38^ The prevalence of lesions with a paramagnetic rim in our study was slightly higher (80 out of 607 lesions, or 13%) compared to previously reported findings (typically ranging from 4.5% to 11%).^12,22,39,40^ This difference could be attributed to our deliberate exclusion of smaller (<100mm^3^) lesions, which are more likely to lack a paramagnetic rim.^4,27^

Our result also confirms a higher proportion of younger patients amongst those displaying PRLs, which is consistent with earlier reports.^14,15,26^

### Relationship between PRL and disease progression

We found that the mere presence of PRLs was not linked to either clinical or radiological measures of disease progression. However, the number of PRL counted in individual patients did show a significant association with measures of total brain atrophy, white matter atrophy and central brain atrophy. Similarly, the volume of PRL lesions significantly correlated with total and central brain atrophy. Furthermore, a significant correlation was observed between PRL volume and change in EDSS during the follow-up period.

Results from earlier studies on the link between paramagnetic rim lesions (PRLs) and disease progression in MS show mixed outcomes. Some studies found a weak to moderate correlation between the presence ^14,38^ or number^18,21,37^ of PRLs and progression in EDSS scores or brain atrophy, ^20^ while others noted that the volume of rim lesions is a more critical factor.^12^ Conversely, some research points to the volume of non-rim lesions as a better predictor of EDSS changes. ^13^ There is also inconsistency in the predictive power of PRL lesions on EDSS vs brain atrophy measures of disease progression. For instance, while ring-positive lesions at baseline predicted EDSS progression, they did not predict the degree of brain atrophy during follow-up^18^ or *vice versa*.^20^ Despite variations due to small sample sizes, differing MRI techniques, and identification methods, there is a general agreement that PRLs tend to correlate with disease progression, supporting our findings.

It is also worth noting that in a number of these studies, the PRL group had significantly (3-10 times) higher baseline lesion volumes,^12,14,18,20,37^ as well as a higher relapse rate (indicative of more active disease)^14^ or more advanced disease at baseline, as measured by EDSS.^20,37^ While our study found that the total lesion load was similar between the PRL and non-PRL groups, it was strongly correlated with the volume of PRLs - a pattern that was also observed between total lesion count and PRL lesion count. Therefore, it is plausible that high T2 lesion volume by itself may drive, at least to some extent, the acceleration of disease progression.

The loss of association between PRL and disability, when adjusted for baseline T2 lesion volume, as demonstrated by Reeves et al.,^14^ also supports this view.

### Are paramagnetic rims present in all new lesions?

We found that every single on-study lesions (i.e., all 32 lesions that developed during the 5-year follow-up period) displayed a paramagnetic rim, which was somewhat unexpected. To ensure the reliability of this result, validation was conducted using two entirely independent cohorts. It confirmed the initial findings, showing that all new lesions larger than 100 mm³ also demonstrated the presence of a paramagnetic rim.

While it is generally accepted that PRLs can arise at the time of lesion formation,^8^ the exact proportion of new lesions that develop into PRLs remains a subject of debate.

Several studies have reported a significant fraction of new or recent MS lesions displaying a paramagnetic rim, while others have not observed this phenomenon. Zhang et al.^27^ identified 50% of new contrast-enhancing lesions (CELs) as rim-positive. Notably, rim-positive lesions were significantly larger than rim-negative lesions at the time of enhancement and remained on average over three times larger than rim-negative lesions, the majority of which did not exceed 100 mm³ at follow-up. Clark et al.^24^ reported that 40% of CELs developed into PRLs. Again, the average lesion volume of PRLs was more than three times larger compared to non-PRL and lesion volume was the main factor predicting the development of a paramagnetic rim. Absinta et al.,^26^ observed a persistent paramagnetic rim in 29% of CELs. The average size of PRL-positive lesions exceeded that of PRL-negative lesions by more than four times. Notably, all but one rim-negative lesion had a volume smaller than 100 mm³, with a significant number not exceeding 10 mm³, suggesting a strong association between lesion size and the presence of a paramagnetic rim. In this study lesion volume at baseline was again the strongest predictor of the persistence of a phase rim at 12 months. In study by Wenzel et al.^23^ 18% of CELs converted into PRLs. Consistent with the studies described above, CELs that converted into PRLs were significantly larger than those that did not.

These findings consistently highlight the critical role of lesion size in the development of paramagnetic rims. All referenced above studies have shown that new lesions exceeding a certain size threshold commonly convert to PRLs. Given our deliberate exclusion of small (<100mm³) lesions in the current study, our results largely align with these observations, further emphasising lesion size as a key determinant in the formation of paramagnetic rims.

Conversely, two studies did not observe paramagnetic rims in any of the newly appeared lesions. ^4,12^ Neither study, however, provided volumetric measurements of the new lesions, which makes it challenging to fully interpret this specific aspect of their findings.

A potential explanation for the link between lesion size and the presence of a paramagnetic rim, first proposed by Absinta et al.,^26^ suggests that the magnitude of the initial inflammation (indicated by the larger size of acute lesions), combined with patient-specific factors such as age, tissue energetics and vascular status triggers a more robust immune reaction at the lesion edge. This immune response aims to isolate the lesion from the surrounding unaffected parenchyma and involves the subtle opening of the blood-brain barrier in capillaries at the lesion edge. This phenomenon, manifesting as a centripetal pattern of contrast enhancement, is associated with the infiltration of blood-derived monocytes/macrophages and the activation of resident microglia,^21^ which is reflected in the appearance of a phase rim. In contrast, smaller lesions experience less intense inflammation and are more conducive to “repair-promoting” mechanisms, such as remyelination and new synapse sprouting^41^. These mechanisms could prevent a significant immune reaction at the lesion edge and reduce the iron accumulation. Additionally, it has been shown that detecting PRLs requires a certain concentration of iron within cells, which may not be reached in smaller lesions.^4^ Therefore, the smaller lesion size reduces the likelihood of paramagnetic rim formation or detection.

Once acute inflammation subsides, new lesions may evolve into chronic active lesions, chronic inactive lesions, or undergo remyelination.^8,42^ However, our results, along with findings from other studies, suggest that the presence of a paramagnetic rim is primarily related to lesion age, manifesting in newly formed lesions that exceed a specific size threshold, regardless of ongoing chronic inflammation activity. This notion is further supported by recent findings from two independent research groups, both of which reported minimal overlap between slowly expanding chronic lesions (SEL) and PRL-positive lesions.^28^,^29^ The association of PRLs with newly or recently formed lesions also helps explain why the joint occurrence of SELs and PRLs found in these studies is linked to greater progression compared to either SELs or PRLs in isolation, since, as we previously demonstrated, both acute and chronic lesional activities independently contribute to the accumulation of tissue damage. ^43^ ^44^

The presence of a paramagnetic rim in new or recent lesions may also explain the reported associations between the presence, number, or volume of PRLs and various measures of disease progression observed in this and other studies. This association likely arises because clinical progression and brain atrophy are more pronounced in patients with recently developed lesions, where the inflammatory process remains active and contributes to ongoing tissue damage.^45^ This is further supported by the younger age of patients with PRLs. Consequently, PRLs could serve as an early indicator of more aggressive disease activity, underscoring the importance of timely detection and targeted therapeutic interventions

The lesion age-dependent nature of the paramagnetic rim is also supported by its evolution over time. Studies have observed that the rim gradually fades,^46^ with reports suggesting a median disappearance time of approximately 7 years.^47^ This is consistent with our findings, which show evidence of rim fading and a significant reduction in the proportion of PRL-positive lesions as lesion age surpasses 5 years.

A fundamental question for future studies is whether all rim-positive lesions gradually evolve into non-rim lesions as they lose the iron deposited during the acute stage of inflammation - given that the paramagnetic properties of MS lesions decay over several years^27^ - or whether the rim persists in CAL, where iron continues to accumulate due to ongoing activity of permanently activated microglia. Answering this question could have significant implications for the clinical use of PRLs. If the former assumption is true, PRLs may not be a suitable biomarker for CAL. Conversely, if the latter assumption is correct, it could also complicate the use of PRLs to detect CAL in cross-sectional or short-term longitudinal studies, including clinical trials, as it may lead to confusion between true chronic active lesions and recently formed lesions.

### Study Limitations

Several study limitations must be acknowledged.

First, the relatively small sample size of 60 patients in the main cohort, while sufficient for detecting broad trends, may limit the generalizability of our findings. Additionally, the exclusion of smaller lesions (<100 mm³) from our analysis could introduce bias, as these lesions might behave differently than larger ones, potentially limiting our understanding of the full spectrum of PRL formation and its implications for disease progression.

Moreover, the study’s reliance on a 5-year follow-up period, while valuable, may not be sufficient to fully capture the long-term impact of PRLs on brain atrophy and clinical outcomes. The inability to accurately determine the exact age of lesions older than 5 years further complicates our analysis, introducing a degree of uncertainty.

It is also possible that other confounding factors, such as variations in treatment adherence or genetic predispositions, which were not fully accounted for in this study, may have influenced our results.

Finally, while our study highlights the association between PRLs and lesion characteristics, the temporal dynamics of PRLs remain incompletely understood. The observed fading of PRLs over time suggests that they may not be a stable marker of chronic active lesions, potentially complicating their use in longitudinal studies or clinical trials aimed at assessing treatment efficacy.

We acknowledge that different treatment options may exert varying effects on the appearance and duration of the paramagnetic rim. Therefore, heterogeneity of treatment at baseline in our study and changes in medication during the follow-up period limit our ability to comprehensively assess their impact on our observations.

In conclusion, our findings indicate that the presence of a paramagnetic rim in MS lesions is a characteristic feature of all newly formed lesions that exceed a specific size threshold, and that this rim gradually diminishes as the lesion ages. As such, this study enhances the understanding of lesion formation and evolution and may have significant implications for using PRLs as a biomarker of lesion activity.

### Data availability

The data that support the findings of this study are available from the corresponding author, upon reasonable request.

## Supporting information

All Supp Data

