## Supplementary material for "The Presence of a Paramagnetic Phase Rim is Linked to Lesion Age in Multiple Sclerosis": All Supp Data

**Sydney cohort (Main study)**

1. **Specific MRI parameters and image processing.**

The following MRI sequences were acquired:

a. Pre- and post-contrast (gadolinium) Sagittal 3D T1: GE BRAVO sequence, duration 4 min each, FOV 256mm, Slice thickness 1mm, TE 2.7ms, TR 7.2ms, Flip angle 12°, Pixel spacing 1mm. Acquisition Matrix (Freq x Phase) is 256x256, which results in 1mm isotropic acquisition voxel size. The reconstruction matrix is 256x256.
b. FLAIR CUBE; GE CUBE T2 FLAIR sequence, duration 6 min, FOV 240mm, Slice thickness 1.2 mm, Acquisition Matrix (Freq x Phase) 256x244, TE 163ms, TR 8000ms, Flip angle 90°, Pixel spacing 0.47 mm. The reconstruction matrix is 512x512.
c. Echo-Planar Imaging based diffusion weighted MRI, duration 9 min (64-directions with 2 mm isotropic acquisition matrix, TR/TE = 8325/86 ms, b = 1000 s/mm^2^, number of b0s = 2).
d. Susceptibility Weighted Angiography (SWAN) sequence, duration 5 min, FOV 230mm, Slice thickness 1mm, Acquisition Matrix (Freq x Phase) 320x224, TE 23.23ms, TR 37.4ms, Flip angle 15°, Pixel bandwidth 244.141 Hz/pixel, Imaging frequency 127.769234 MHz, Echo Train Length 6, Number of Averages 0.701292, and Specific Absorption Rate (SAR) 0.168281 W/kg. Spacing between slices is 1mm.

**MRI image pre-processing:**

The baseline T1-weighted imaging was realigned to Anterior and Posterior Commissure (AC-PC) orientation. Using FLIRT (FSL, FMRIB Software Library), follow-up T1 images were co-registered to initial (month 0) AC-PC space by applying transformation matrices derived from linear co-registration between baseline AC-PC aligned brain and follow-up native T1 brain images. In parallel, diffusion MRI was corrected for motion and eddy-current distortion in FSL, then EPI susceptibility distortion was minimized by applying deformation maps generated from nonlinear co-registration between DWI b0 brain images and T1-weighted images at each time-point using ANTS (Advanced Normalization Tools). Subsequently, tensor reconstruction was performed in MRtrix3. Tensor and FLAIR images were then linearly co-registered to corresponding T1 AC-PC images at each time point. Finally, SWAN images were linearly co-registered to the corresponding T1 AC-PC images, and the transformation was applied to the PHASE image to align all the images within the same space.

Phase images generated from SWAN were then analysed alongside corresponding FLAIR sequences to identify PRL.

The lesion segmentation was performed on unprocessed T1-weighted and FLAIR images. Following segmentation, lesion masks were transformed to ACPC space through linear registration to ensure consistent spatial alignment across scans.

All reported T1 intensity values were WhiteStripe-normalized, ensuring that the intensity values are scaled relative to the reference white matter intensity.^32^ This method involves identifying a stable region of white matter intensities, referred to as the "white stripe," which serves as a reference for intensity scaling.

### ***Validation datasets. Review!!!***

To validate the relationship between the presence of a paramagnetic rim and lesion age, data from additional cohorts were used, including 100 RRMS patients from Charité-Universitätsmedizin Berlin, Germany, and 163 CIS and MS patients from Vall d’Hebron University Hospital, Barcelona, Spain. Each patient underwent at least 3 consecutive MRI scans. The average time of follow-up was 43 ±23 months for the Berlin cohort and 43 ± 19 months for the Barcelona cohort. Similar to the main study, PRLs were assessed at the final time point.

Using T2-weighted FLAIR and filtered phase images (co-registered to T1-weighted images) a minimum of three time points were reviewed. First, new lesions ≥100mm^3^ on the intermediate FLAIR scan (compared to baseline) were identified, and their presence was confirmed on the latest FLAIR image. Each new lesion was then assessed on the last available filtered phase image for the presence of a paramagnetic rim, as done for the original study cohort. The Berlin images were reviewed by AK and TU and the Barcelona images were reviewed by AK and MC. Only lesions in susceptibility visible areas were analysed (i.e. areas free of potential susceptibility distortion such as infratentorial area and cortex and deep grey matter regions). Any discrepancies were resolved by consensus.

While the main study was conducted on a GE scanner, which produces a 'right-handed' phase map, our validation dataset was acquired on a Siemens scanner with a 'left-handed' phase map. Consequently, these phase orientation differences may result in distinct presentations of hypointense and hyperintense rims. This factor was carefully considered when detecting PRLs to maintain consistency in analysis across systems.

**DTI data processing:**

Diffusion weighted MRI data (dMRI) were pre-processed using tools provided by the software suites MRtrix3, FSL, and ANTs. Specifically, dMRI data were first denoised, then potential Gibbs-ringing artefacts were removed. The dMRI data were then corrected for bias field inhomogeneities using the ANTs N4 algorithm. a dMRI brain mask was then estimated using BET and used as an input, alongside the dMRI data in order to correct for subject movement in the acquisition. Finally, phase distortion correction was applied using a non-linear registration method outlined below.

To correct for phase distortion within the dMRI data, first the brain was segmented from the corresponding T1w dataset, and a single B0 volume was extracted from the dMRI dataset. The T1w brain was then used as a mask to invert the contrast of the T1w image. A rigid-body registration was then performed on the inverted T1w image with the B0 volume as a target, which aligned the two images spatially. Non-linear registration was then performed using ANTs. The registration steps were comprised of a rigid body, then affine, and then SyN registration algorithm. The transformations and warps calculated from the non-linear registration steps were then applied to the entire dMRI dataset to correct for phase distortion artefacts.

**The criteria for PRLs**

The criteria for PRLs included a hypointense paramagnetic rim that is continuous along at least two-thirds of the outer lesion edge on the slice of maximum visibility and is visible on at least 2 consecutive slices, the rim co-localises with the edge of all or part of the lesion core that is hyperintense on FLAIR image and non-enhancing on post-contrast T1 sequence.

**Disease-modifying therapy**

Disease-modifying therapy categories for main cohort were grouped into three categories: no treatment, moderate-efficacy treatment (e.g., interferon, glatiramer acetate, teriflunomide, fingolimod and dimethyl fumarate) and high-efficacy treatment (e.g., natalizumab, alemtuzumab, ocrelizumab).^34, 35^ At the start of the study, 10 patients were on moderate-efficacy treatment, while 42 patients were receiving high-efficacy therapies, and 8 patients were not receiving any treatment. Throughout the study, 3 patients started treatment, 2 patients stopped treatment, and 5 patients transitioned between treatment categories. By the end of the study period, 8 patients remained on moderate-efficacy treatment, and 45 patients were on high-efficacy therapies. Treatment modifications were made by the treating physician based on clinical and radiological activity, with 50 patients remaining in their initial treatment category for the duration of the study.

Supplementary Table 1.

The Berlin cohort was scanned using a 3T Siemens Prisma scanner equipped with a 64-channel head coil. People with RRMS and a minimum of 3 time-points were included (minimum time between time-points 6 months).

The Barcelona cohort was scanned using a 3T Siemens Magnetom Trio scanner with a 12-channel phased array head coil. People with CIS/RRMS and 3 time-points were included (baseline, 1 and 3 years).

| Cohort | Imaging Sequence | Image resolution (mm3) |
| --- | --- | --- |
| Berlin | 3D T1-weighted MPRAGE | 0.8 x 0.8 x 0.8 |
|  | T2-weighted FLAIR | 0.8 x 0.8 x 0.8 |
|  | 3D GRE SWI with filtered phase | 0.7 x 0.7 x 2 |
| Barcelona | 2D T1-weighted | 1 x 1 x 1 |
|  | 2D/3D T2-weighted FLAIR | 0.5 x 0.5 x 3 /  1 x 1 x 3 |
|  | 2D GRE SWI with filtered phase | 0.5 x 0.5 x 3 |

Figures

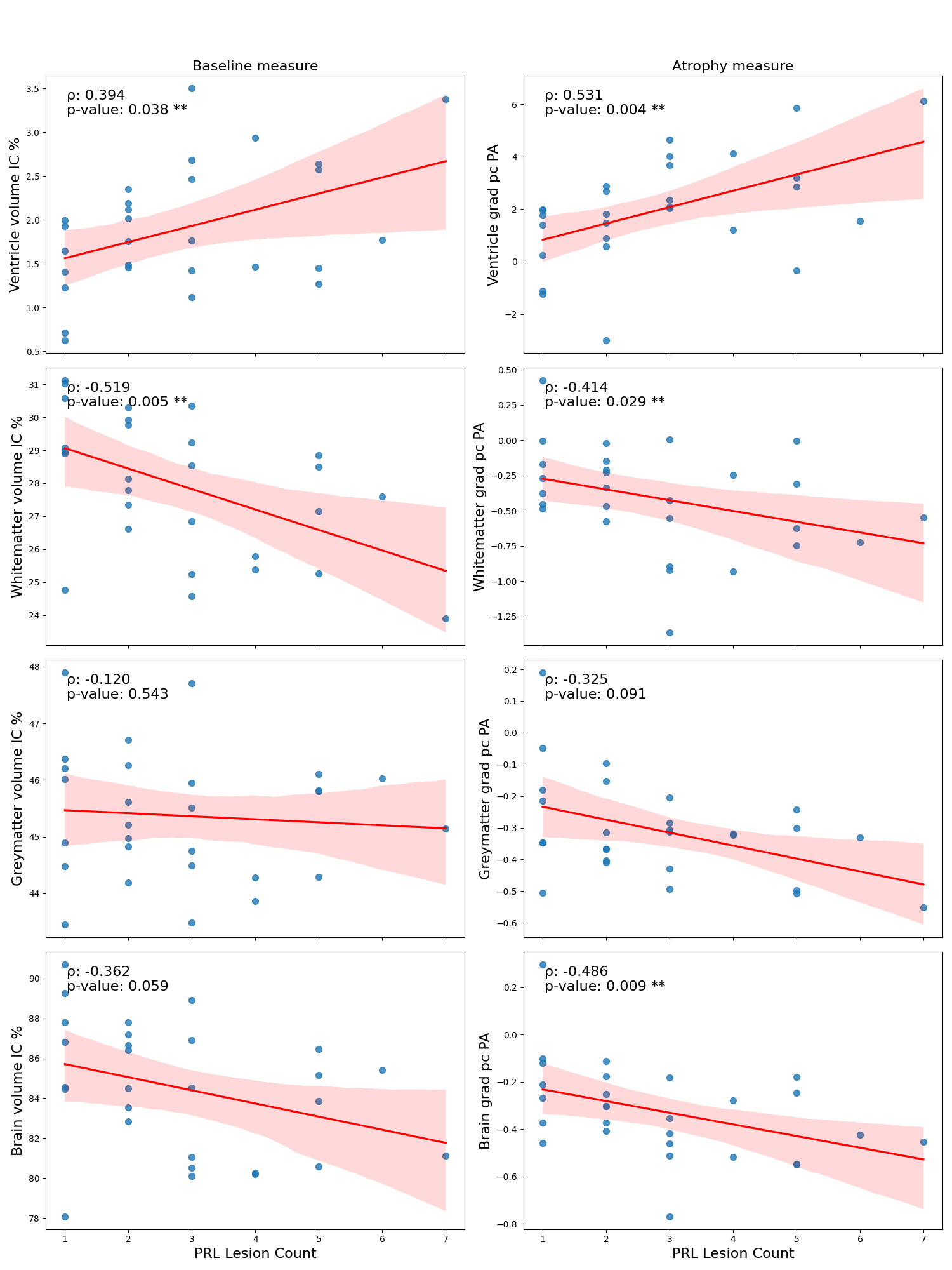

**Suppl. Figure 1:** Correlation between radiological measures and PRL count. The first column shows PRL count versus baseline radiological measures at the SWAN timepoint, and the second column shows PRL count versus atrophy measures. Correlations were assessed using Spearman’s rank, with rho and p values displayed for each plot. Red lines represent the line of best fit, and the shaded areas around them indicate the 95% confidence intervals.
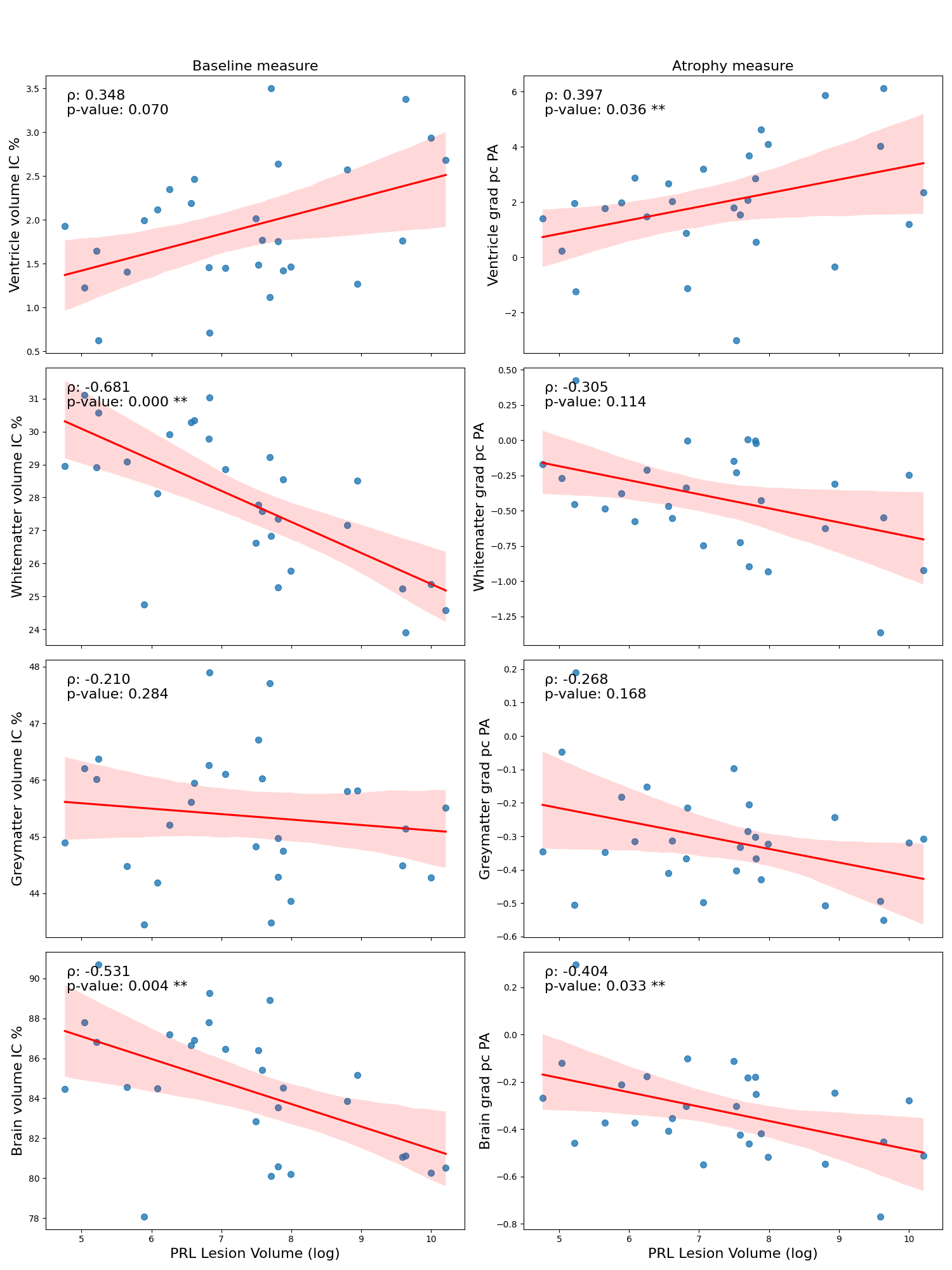

**Suppl Figure 2: Correlation between radiological measures and PRL volume.** The first column shows PRL volume versus baseline radiological measures at the SWAN timepoint, and the second column shows PRL volume versus atrophy measures. Note that PRL volume was log-transformed for visualisation purposes (this does not affect the correlation or p-values). Correlations were assessed using Spearman’s rank, with rho and p values displayed for each plot. Red lines represent the line of best fit, and the shaded areas around them indicate the 95% confidence intervals.
